# High- Versus Low-Dose Dexamethasone for the Treatment of COVID-19-related Acute Respiratory Distress Syndrome: A Multicenter and Randomized Open-label Clinical Trial

**DOI:** 10.1101/2021.09.15.21263597

**Authors:** Luis Patricio Maskin, Ignacio Bonelli, Gabriel Leonardo Olarte, Fernando Palizas, Agostina E Velo, María Fernanda Lurbet, Pablo Lovazzano, Sophia Kotsias, Shiry Attie, Ignacio Lopez Saubidet, Natalio D Baredes, Mariano Setten, Pablo Oscar Rodriguez

**Author notes:** Corresponding Author: Pablo O. Rodriguez, Unidad de Terapia Intensiva, Hospital Universitario Sede Pombo, CEMIC (Centro de Educación Médica e Investigaciones Clínicas), Av. Cnel. Diaz 2423, Zip Code: 1425, Ciudad Autónoma de Buenos Aires, Argentina.

## Abstract

**Purpose:** To determine whether high-dose dexamethasone increases the number of ventilator-free days (VFD) among patients with acute respiratory distress syndrome due to coronavirus disease 2019 (C-ARDS)

**Materials:** A multicenter randomized controlled trial in adults with C-ARDS. Patients received 16 mg/d of dexamethasone intravenously for five days followed by 8 mg/d of dexamethasone for five days, or 6 mg/d of dexamethasone intravenously for 10 days.

**Results:** Data from 98 patients who received at least one dose of dexamethasone were analyzed. At 28 days after randomization, there was no difference between high- and low-dose dexamethasone groups in VFD (median, 0 d [interquartile range (IQR) 0–14 d] versus 0 d [IQR 0–1 d]; *P*=0.231). The cumulative hazard of successful discontinuation from mechanical ventilation was increased by the high-dose treatment (adjusted sub-distribution hazard ratio: 1.84; 95% CI: 1.31 to 2.5; *P*<0.001). None of the prespecified secondary and safety outcomes showed a significant difference between treatment arms.

**Conclusions:** Among patients with C-ARDS, the use of higher doses of dexamethasone compared with the recommended low-dose treatment did not show an increase in VFD. However, the higher dose significantly improved the time required to liberate them from the ventilator

Clinical Trial Registered: NCT04395105

## INTRODUCTION

In December 2019, an outbreak of coronavirus disease 2019 (COVID-19), which was caused by severe acute respiratory syndrome coronavirus 2 (SARS-CoV-2), broke out in Wuhan, China^1,2^. The World Health Organization (WHO) declared it a significant threat to international health. COVID-19 mainly affected the respiratory system with some patients rapidly progressing to acute respiratory distress syndrome (ARDS). Many of them will require mechanical ventilation for a long time, overcrowding the health system. The COVID-19-related ARDS (C-ARDS) main pathological pattern is diffuse alveolar damage^3^, and there is evidence that the dysregulated inflammatory response may worsen the prognosis.

Corticosteroids might exert an effect in controlling this exacerbated response. Over the last decades, many clinical studies have tested the utility of corticosteroids in critically ill patients with ARDS having inconsistent findings^4–6^. Nonetheless, several trials evaluated the role of corticosteroids for ARDS treatment in non–COVID-19^7,8^ and COVID-19 patients,^9–12^ suggesting a decrease in 28-day mortality in patients with oxygen needs or mechanical ventilation. Although the benefit was considered a general class effect of glucocorticoids, various dose regimens were used, leaving the question of a dose–effectiveness relationship less definitively answered^13^.

The dexamethasone for COVID-19-related ARDS randomized clinical trial was conducted to evaluate the effectiveness and safety of high-versus low-dose dexamethasone in patients with C-ARDS. The hypothesis was that high-dose dexamethasone would increase the number of ventilator-free days (VFD) during the first 28 days.

## METHODS

### Study Design and Oversight

We conducted an investigator-initiated, multicenter, randomized, open-label clinical trial in four intensive care units (ICUs) in Argentina. The trial protocol and the statistical analysis plan had been previously published^14^ (Supplement 1). The study was approved at the Research Ethics Committee of CEMIC (Buenos Aires, Argentina) and by the Argentine Society of Intensive Care Medicine (SATI) Ethics Committee. Either a written or an electronic informed consent (REDCap electronic consent framework) was obtained from the legal representative of each patient. The study was registered in PRIISA.BA (code 1264) and in ClinicalTrials.gov (NCT04395105).

### Patients

We enrolled patients aged 18 years old or more, who had ARDS according to the Berlin Definition criteria^15^, who had confirmed SARS-CoV-2 infection by reverse transcription polymerase chain reaction and were receiving mechanical ventilation for less than 72 hours. The exclusion criteria were pregnant or breastfeeding women, terminal disease, therapeutic limitation, severe immunosuppression, chronic treatment with glucocorticoids, participation in another randomized clinical trial, prior use of dexamethasone for COVID-19 (> 5 days), or consent refusal (e-Methods Supplement 2).

### Trial Procedures

Treatment allocation was performed through an online web-based system (REDCap) ^16^ using a permuted random block sequence stratified by center.

The study was originally designed before RECOVERY trial publication, and the control group had not included corticosteroids^9^. Soon after the pre-publications of these results, we amended the protocol to include a low-dose dexamethasone in the control arm. Thereafter, eligible patients were randomly assigned in a 1:1 ratio to receive high- or low-dose dexamethasone plus standard care. The former was 16 mg dexamethasone administered intravenously once daily for five days, followed by 8 mg intravenously administered once daily for additional five days. The low-dose group received 6 mg of dexamethasone per day for 10 days according to the RECOVERY trial. The investigators who assessed the outcomes were not blinded for the assigned treatment.

Standard care was not regulated by the protocol. Nonetheless, it was suggested to treat the patients according to their institutional protocols or the international guidelines for ARDS^17^, antibiotics, and hemodynamic support for COVID-19 infection^6,18^. The ventilator liberation protocol was defined by each site. Nevertheless, it was recommended to daily evaluate the eligibility of the patients to perform a spontaneous breathing trial^17^.

### Clinical and Laboratory Data

Data on demographic characteristics, physiological variables, severity scores, timing from ARDS diagnosis to randomization, corticosteroid use, COVID-19 therapies, and other clinical and laboratory data were collected. The use of sedatives, neuromuscular blocking agents, prone positioning, vasopressors, renal replacement therapy, and extracorporeal membrane oxygenation (ECMO) were registered daily throughout the first 28 days since randomization or until ICU discharge. For that period, we collected information on the use of mechanical ventilation, respiratory monitoring, and other oxygen supportive therapies. Data regarding infections, glycemic control, muscle dysfunction, and delirium were also collected as a safety measurement (e-Methods Supplement 2).

The patients were followed up for 28 days after randomization or until hospital discharge, whichever occurred first. The vital status was assessed 28 and 90 days after randomization when needed by a phone interview.

### Outcomes

The primary outcomes were VFD during the first 28 days, defined as the number of days alive and free from mechanical ventilation up to the 28th day from randomization. For the patients who died, the number of VFD was set as 0. As a co-primary outcome, the time to complete and successful discontinuation of mechanical ventilation or death was calculated from randomization. The former was defined as the difference in time between randomization and the last day spent on mechanical ventilation without further invasive respiratory support^18^.

The secondary outcomes were all-cause mortality at 28 and 90 days, the rate of nosocomial infections, the daily value of glucose and insulin dose, muscle strength score, and the frequency of delirium within 28 days of randomization (e-Methods Supplement 2).

### Statistical Analysis

No reliable data on ARDS caused by COVID-19 were available at the trial design to allow for an accurate sample size calculation. Therefore, we employed the data from a recently published multicenter randomized trial of non–COVID-19 ARDS^7^ for our sample size calculation. The sample size was calculated at 142 patients in each group to detect a difference of three VFD between groups, assuming a mean and a standard deviation of 9 days with a two-sided α level of 0.05 and a power of 80%.

The quantitative variables were expressed as mean and standard deviation or median (25th to 75th percentile range) for normal or non-normal data distribution assessed with the Shapiro–Wilks normality test. The comparison of these variables between experimental treatments was performed using a t-test or a Wilcoxon rank sum test. Furthermore, the proportions were compared with the Fisher exact or chi-squared tests.

The main outcome variable analysis (VFD at day 28) was compared between treatments as stated above. As the Wilcoxon rank sum test does not provide a measure of effect, we calculated the basic bootstrap 95% confidence interval of the difference between treatment arm medians as an exploratory analysis^19^. To further explore the potential effect of the treatments, a time-to-event analysis for competing risks was performed to evaluate the time to the complete and successful discontinuation of mechanical ventilation within 28 days. In this analysis, death was considered the competing event, and cumulative incidence curves according to the treatment allocations were constructed. Furthermore, a competing-risks regression model for clustered data was utilized to estimate the effect of treatment on the sub-distribution hazard adjusted for APACHE II and ARDS severity^20^. Each ICU was included as a cluster in the model^19^.

The probability of survival at 90 days was evaluated with a Kaplan–Meier analysis, and the log-rank test was used for comparison between treatments. A Cox proportional hazards regression model was fit to adjust the treatment effect with APACHE II and ARDS severity.

The rate of infection observed within 28 days of inclusion was calculated using a Poisson regression model, and the experimental treatment was used as a predictor. Additionally, the incidence rate ratio for high-dose dexamethasone treatment and its 95% confidence interval were calculated. Mixed effects linear models were also employed to evaluate the interaction between the treatment allocation and the time after the inclusion of glucose blood levels and insulin doses. To avoid pseudo-replications, each subject was used as a random effect.

A modified intention-to-treat approach was used for the analysis, including only data from those patients who received at least one dose of dexamethasone after inclusion. A two-sided *P* value of less than .05 was considered statistically significant, and all the analyses were performed using R software version 3.6.1^21^.

### Premature Trial Termination

During the recruitment period, the use of dexamethasone in the early course of COVID-19 was widely recommended by several trials^9–12^, which implied that an increasing number of patients were admitted to the ICU with complete corticosteroid treatment. Moreover, the use of high-flow nasal cannula oxygen therapy for severe COVID-19 pneumonia was encouraged^6,22^, thereby delaying the initiation of mechanical ventilation in some patients. Therefore, the recruitment rate lowered substantially. By the end of March 2021, we estimated that the time required to achieve the calculated sample size would be greater than three years (e-Figure 1 and e-Figure 2 Supplement 2). As COVID-19 management is rapidly evolving, the research question would probably be obsolete at the end of this time. Due to this estimation, the investigators decided to prematurely terminate the trial on April 5, 2021. No interim data analysis of efficacy or safety was performed before this decision.

## RESULTS

### Patients

Between June 17, 2020, and March 27, 2021, 211 C-ARDS patients were screened. One hundred were enrolled, of whom 49 were randomized to the high-dose dexamethasone and 51 to the control group (Figure 1). The data from the first two participants from the control group were not analyzed as they did not receive any dexamethasone. The baseline characteristics were well balanced between groups (Table 1), except for APACHE II and time in mechanical ventilation before inclusion. At randomization, ventilator settings, respiratory system mechanics, and gas exchange parameters were not different between the treatment groups (Table 1). The baseline laboratories and additional treatments did not differ between groups (e-Table 1 and e-Table 2 Supplement 2).

**Figure 1.**
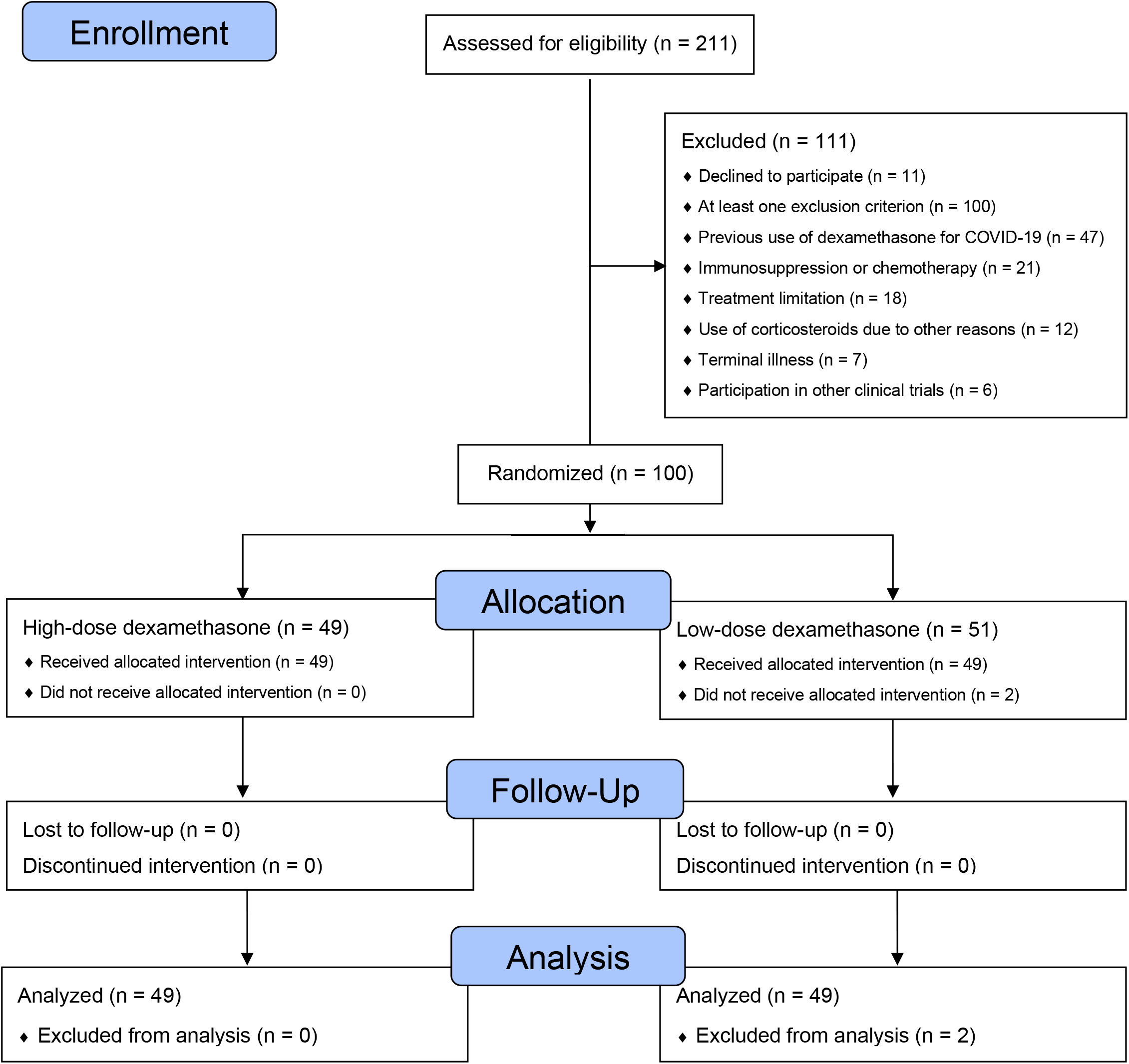
Flow diagram of the study. Abbreviations: COVID-19, Coronavirus disease 2019

**Table 1.** Baseline patients’ characteristics^a^

|  | Low-dose<br>dexamethasone<br>*n*=49 | High-dose<br>dexamethasone<br>*n*=49 | All<br>patients<br>*n*=98 |
| --- | --- | --- | --- |
| Age (y) | 60.04±13.08 | 63.57±13.59 | 61.81±13.38 |
| Gender (female) | 16 (33%) | 13 (26%) | 29 (30%) |
| Weight (kg) | 97 (80–110) | 92 (80–110) | 95 (80–110) |
| BMI (kg/cm<sup>2</sup>) | 33.8 (28.3–38.9) | 31.1 (28.3–38.1) | 32.7 (28.3–38.7) |
| APACHE II <sup>b</sup> | 12.1±4.6 | 14.7±5.5 | 13.4±5.2 |
| SOFA<sup>c</sup> | 6 (3–7) | 5 (3–6) | 6 (3–7) |
| Charlson's comorbidity index<sup>d</sup> | 0 (0–1) | 1 (0–1) | 0 (0–1) |
| Symptoms (days from onset) | 10 (8–13) | 9 (7–12) | 10 (7–13) |
| Hospital length of stay (days) | 3 (2–6) | 4 (2–8) | 4 (2–7) |
| Length of MV (hours) | 22 (10–31) | 31 (18–49) | 25.5 (13–44) |
| *ARDS severity* |  |  |  |
| Mild | 1 (2%) | 4 (8%) | 5 (5%) |
| Moderate | 30 (61%) | 30 (61%) | 60 (61%) |
| Severe | 18 (37%) | 15 (31%) | 33 (34%) |
| *Ventilator setting and monitoring* |  |  |  |
| PEEP (cmH<sub>2</sub>O) | 13.1±2.9 | 12.7±2.6 | 12.9±2.8 |
| Tidal volume (mL/kg) | 6.3 (6–6.9) | 6.3 (6–6.9) | 6.3 (6–6.9) |
| Respiratory rate (bpm) | 25 (24–27) | 25 (24–27) | 25 (24–27) |
| Plateau pressure (cmH<sub>2</sub>O) | 24 (22–27) | 24 (22–27) | 24 (22–27) |
| Driving pressure (cmH<sub>2</sub>O) | 11 (10–12) | 11 (9–14) | 11 (10–13) |
| Compliance (mL/cmH<sub>2</sub>O) | 35 (30–40) | 37 (32–45) | 36 (31–42) |
| *Gas exchange* |  |  |  |
| pH | 7.31±0.07 | 7.33±0.06 | 7.32±0.06 |
| PaCO<sub>2</sub> (mmHg) | 47 (41–53) | 47 (40–51) | 47 (41–53) |
| PaO<sub>2</sub>/FiO<sub>2</sub> ratio (mmHg) | 182±55 | 207±73 | 194±66 |
| *Other characteristics* |  |  |  |
| Prone positioning (%) | 12 (24%) | 8 (16%) | 20 (20%) |
| NMBD use (%) | 35 (71%) | 39 (80%) | 74 (76%) |
| Vasoactive drugs (%) | 35 (71%) | 25 (51%) | 60 (61%) |
| Renal replacement (%) | 0 (0%) | 1 (2.04%) | 1 (1%) |

Abbreviations: APACHE, Acute Physiology and Chronic Health Evaluation; ARDS, acute respiratory distress syndrome; BMI, body mass index; MV, mechanical ventilation; NMBD, neuromuscular blockade drugs; PaCO<sub>2</sub>, partial pressure of arterial oxygen; PaO<sub>2</sub>, partial pressure of arterial carbon dioxide; PaO<sub>2</sub>/FiO<sub>2</sub> ratio, partial pressure of arterial oxygen to the fraction of inspired oxygen ratio; PEEP, positive end expiratory pressure; SOFA, Sequential Organ Failure Assessment.
<sup>a</sup> Continuous variables are presented as median (25<sup>th</sup> to 75<sup>th</sup> interquartile range) or mean ± SD.
<sup>b</sup> The Acute Physiology and Chronic Health Evaluation II ranges from 0 to 71, with higher scores indicating a higher risk of death. It is calculated from 14 variables within 24 hours of admission to the intensive care unit.
<sup>c</sup> The Sequential Organ Failure Assessment was measured in six organ systems (cardiovascular, hematologic, gastrointestinal, renal, pulmonary, and neurologic), with each organ having a score from 0 to 4, resulting in an aggregated score that ranges from 0 to 24, with higher scores indicating greater dysfunction.
<sup>d</sup> Charlson's comorbidity index predicts the one-year mortality for a patient who may have a range of comorbid conditions, from a total of 22. Each condition is assigned a score of 1, 2, 3, or 6, depending on the risk of dying associated with each one. Scores are summed to provide a total score to predict mortality.

### Interventions

The durations of dexamethasone treatment were 10 (7–10) and 9 (7–10) days in the high- and low-dose groups, respectively (P =.339). After the intervention phase, 20 (20.4%) patients received corticosteroids, mainly hydrocortisone, due to septic shock (10 in each group, P >.999).

### Primary Outcomes

The VFD within 28 days of the inclusion in the trial was not different between the study groups (Table 2; 0 (0–14) versus 0 (0–1) days for the high- and low-dose dexamethasone groups; P =.231). The difference between these medians was 0 (bootstrap 95% CI: 0 to 2) days. The times spent on mechanical ventilation after randomization were 12 (6–26) versus 19 (9–32) days for the high- and low-dose groups (P =.078). The difference was -7 (bootstrap 95% CI: -17 to 3) days. When only patients discharged alive without mechanical ventilation were considered, these times were 14 (8–26) versus 27 (10–31.5) days for the high- and the low-dose of the dexamethasone groups (P =.154), and the within-median difference was -13 (bootstrap 95% CI: -31 to -6) days.

**Table 2.** Primary and secondary outcomes according to the treatment allocation

|  | Low-dose<br>dexamethasone |  | High-dose<br>dexamethasone |  | All<br>Patients |  | P |
| --- | --- | --- | --- | --- | --- | --- | --- |
|  | Statistic | n | Statistic | n | Statistic | N |  |
| *Primary Outcome* |  |  |  |  |  |  |  |
| 28-VFD (days) | 0 (0–1) | 49 | 0 (0–14) | 49 | 0 (0–8.75) | 98 | .231 |
| *Total time on MV (days)* |  |  |  |  |  |  |  |
| All | 19 (9–32) | 49 | 12 (6–26) | 49 | 15.5 (7–30) | 98 | .078 |
| Discharged without MV | 27 (10–31.5) | 23 | 14 (8–26) | 25 | 18 (8–30) | 58 | .154 |
| *Intensive care unit outcome* |  |  |  |  |  |  |  |
| Mortality (%) | 24 (49%) | 49 | 21 (43%) | 49 | 45 (46%) | 98 | .685 |
| Length of stay (days) |  |  |  |  |  |  |  |
| All | 24 (10–36) | 49 | 15 (9–28) | 49 | 17 (9.25–35) | 98 | .137 |
| Survivors | 33 (16–43) | 25 | 18.5 (13–44) | 28 | 27 (14–43) | 53 | .397 |
| *Hospital outcome* |  |  |  |  |  |  |  |
| Mortality (%) | 24 (49%) | 49 | 22 (45%) | 49 | 46 (47%) | 98 | .840 |
| Length of stay (days) |  |  |  |  |  |  |  |
| All | 25 (11–41) | 49 | 22 (11–43) | 49 | 23.5 (11–42.5) | 98 | .365 |
| Survivors | 36 (25–46) | 25 | 30 (20.5–54) | 27 | 36 (21–52) | 52 | .905 |
| *Mortality rate* |  |  |  |  |  |  |  |
| 28-day (%) | 19 (39%) | 49 | 20 (41%) | 49 | 39 (40%) | 98 | >.999 |
| 90-day (%) | 23 (47%) | 49 | 23 (47%) | 49 | 46 (47%) | 98 | >.999 |

Abbreviations: MV, mechanical ventilation; VFD: ventilator free days

Figure 2 illustrates the cumulative incidence curves for successful discontinuation from mechanical ventilation and death within 28 days according to the treatment group. The unadjusted sub-distribution hazard ratio for the former of high-dose dexamethasone compared with the low-dose was 1.6 (95% CI: 1.1 to 2.33, P =.013). After adjustment with APACHE II and ARDS severity (Table 3), this ratio was 1.84 (95% CI: 1.31 to 2.59, P <.001).

**Figure 2.**
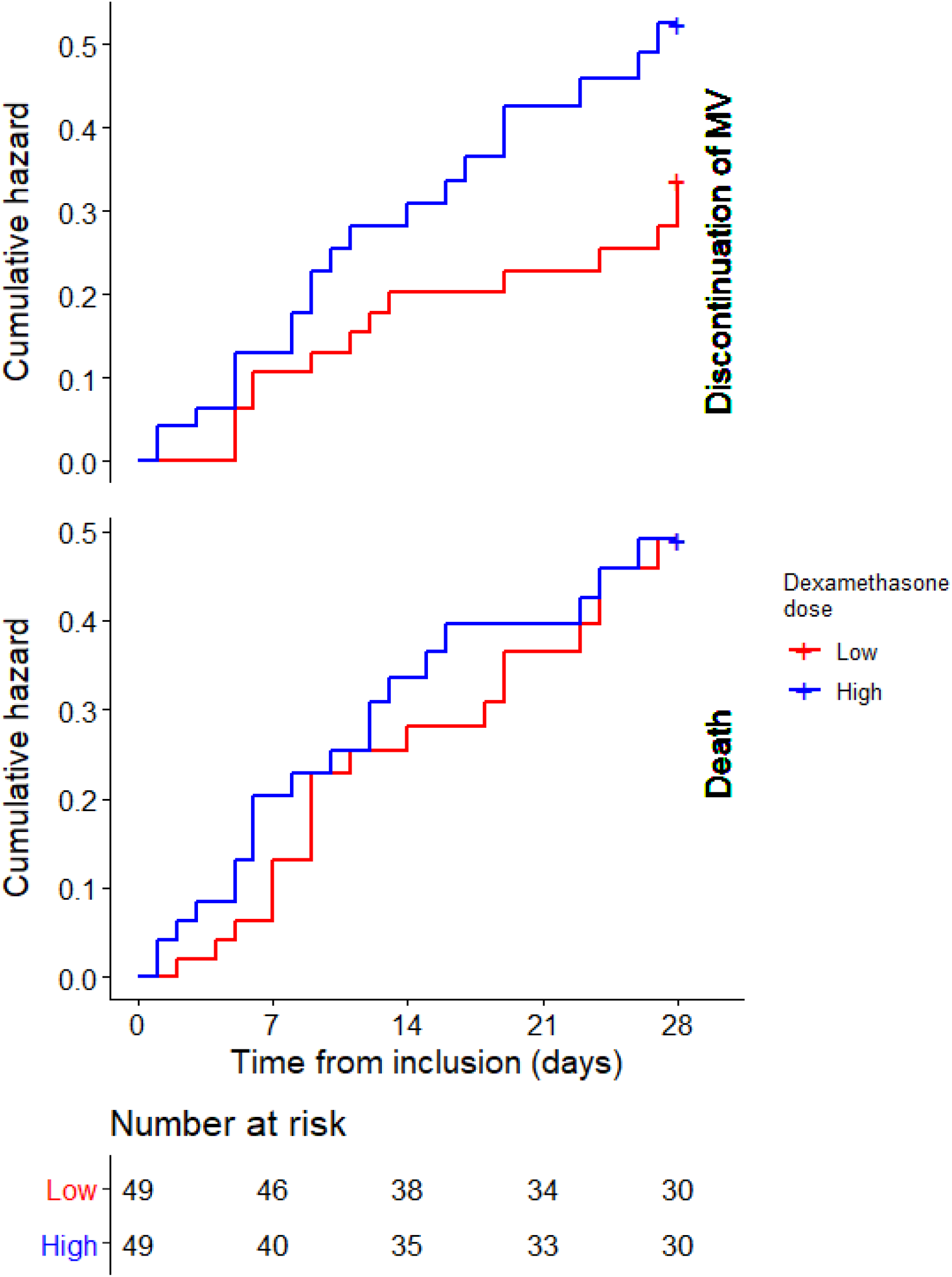
Probability of the successful discontinuation of mechanical ventilation or death within 28 days of the randomization. Cumulative incidence curves of the successful discontinuation of mechanical ventilation (upper panel) and death (lower panel) according to the treatment allocation within 28 days of randomization. The unadjusted sub-distribution hazard ratio for the discontinuation of the mechanical ventilation of high-dose dexamethasone was 1.6 (95% CI: 1.1 to 2.33, P =.013). Abbreviation: MV, mechanical ventilation; CI, confidence interval

**Table 3.**
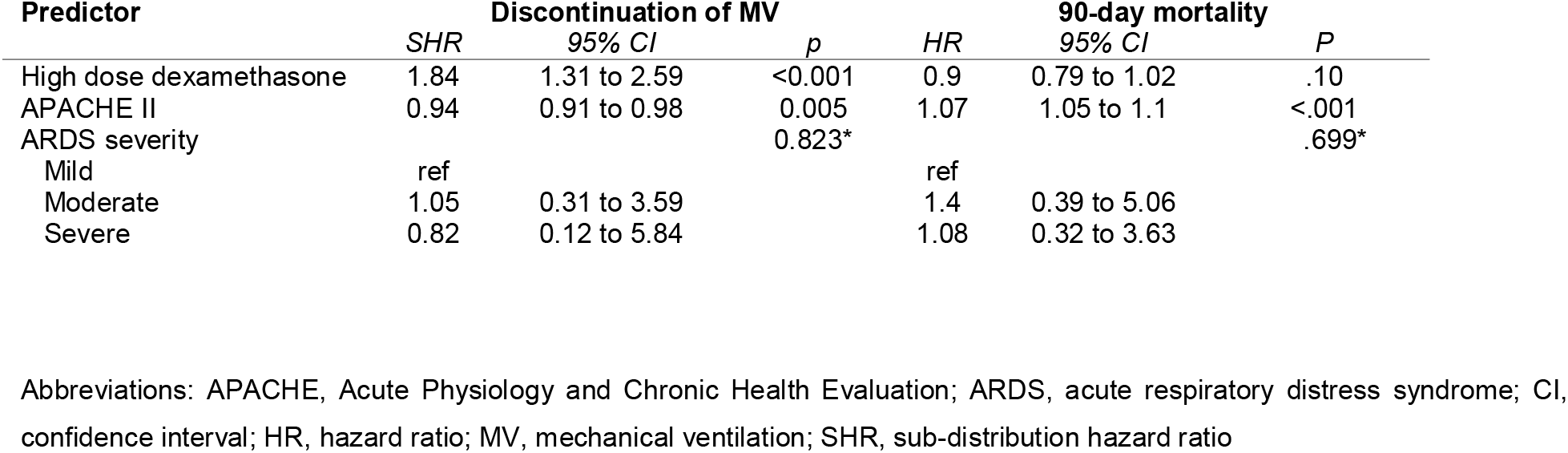
Summaries of the competing-risks regression for time to the successful discontinuation of mechanical ventilation at 28 days and the Cox regression model for 90-day mortality

### Secondary Outcomes

By day 28, 20 (41%) patients in the high-dose dexamethasone group and 19 (39%) in the low-dose group had died (P >.999). The ICU and hospital mortality were also similar between groups. In addition, the length of stay in the ICU in the high-dose dexamethasone group was 15 (9–28) days versus 24 (10–36) days for the low-dose group (P =.137), and the difference between these medians was -9 (bootstrap 95% CI: -20 to 0) days. The hospital length of stay was not affected by the treatment allocation.

Vital status 90 days after randomization was available from all the patients. Forty-six (47%) of them had died, 36 were at home, 11 were in chronic care facilities, and five were still in the hospital. e-Figure 3 shows that the 90-day probability of survival according to treatment allocation was not statistically different (log-rank P =.862). Table 3 exhibits the adjusted hazard ratio of high-dose dexamethasone was 0.9 (95% CI: 0.79 to 1.02, P =.10).

#### Protocol-defined Safety Outcomes

Microbiologically confirmed infections were diagnosed in 72 (73%) patients during the first 28 days. The estimated rate of the infections of the control arm was 2.42 (95% CI: 1.92 to 3) per patient per month, and the incidence rate ratio of the high-dose group was 1.12 (95% CI: 0.81 to 1.54, P =.502).

Both groups had a comparable peak finger-prick glucose and daily insulin use. The patients from both groups had low measurements of muscle strength and frequently experienced delirium. No differences between treatments were seen in these two outcomes (e-Results Supplement 2).

## DISCUSSION

In this multicenter randomized, open-label, clinical trial involving 98 adults mechanically ventilated for C-ARDS, a 10-day course of intravenous high-dose dexamethasone compared with low-dose dexamethasone, both in addition to the standard care, did not show an increase in the VFD during the first 28 days, but significantly increased the hazard of the successful discontinuation of mechanical ventilation. High-dose dexamethasone did not affect the probability of survival at 90 days and was not associated with an increased risk of corticosteroid-associated safety issues.

The RECOVERY trial showed a survival advantage for hospitalized COVID-19 patients with respiratory support treated with 6 mg dexamethasone once daily for up to 10 days. The corticosteroid was administered either orally or intravenously. Although dexamethasone pharmacokinetics has not been studied in critically ill patients, its volume of distribution was approximately 1 liter/kg of body weight in a study involving hospitalized patients due to community acquired pneumonia^23^. Interestingly, this study reported an equivalent area under the curve of serum dexamethasone concentration following a single oral administration of 6 mg or a 4 mg intravenous bolus. As many severe COVID-19 patients are obese or have overt overweight, doses of dexamethasone larger than 6 mg would probably be required in these cases. The initial median dose of intravenous dexamethasone in our patients from the low-dose group was 0.06 (0.05–0.08) mg/kg, which is equivalent to 0.42 (0.36–0.5) mg/kg of prednisone. This dose might be considered low for a critically ill patient suffering from an acute and severe inflammatory lung disease. Conversely, the high-dose dexamethasone arm initially received 0.17 (0.15–0.2) mg/kg of the drug, which is equivalent to 1.16 (0.97–1.33) mg/kg of prednisone.

The primary endpoint of our study was 28-day VFD, which has frequently been utilized as a failure-free outcome in the critical care literature. It is a composite variable, which combines the time required for the liberation of mechanical ventilation and the risk of death. Thus, a certain value could arise from the divergent combinations of its components. When the former is too long or the latter too high, an excess of zeros may preclude the detection of any effect related to the intervention with standard statistics^24^. This explains why the median 28-day VFD of our patients was 0. To overcome these issues, we also decided to use a competing-risks analysis to evaluate the effect of the intervention in our statistical plan. Using this approach, even if we could not attain the planned sample size, a high-dose dexamethasone independently reduced the time required for liberating patients from the ventilator. In fact, in an exploratory comparison of the duration of ventilatory support, this time was 13 (95% CI: 6 to 31) days lower in survivors of the high-dose dexamethasone group. To our knowledge, this is the first study that displays a shorter time of invasive mechanical ventilation with dexamethasone doses higher than those recommended. Tomazzini et al.^10^ reported results from the CoDEX trial, which compared a high-dose dexamethasone versus no corticosteroids in a sample of patients with baseline clinical characteristics like ours. They did not observe a difference between the mean duration of mechanical ventilation in the overall population (12.5 versus 13.9 days). Contrarily, Villar et al.^7^ showed a mean difference of 5.3 days (14.2 ± 13.2 versus 19.5 ± 13.2) in the survivors of ARDS non-related to COVID-19 treated with high dexamethasone versus usual care.

Survival probability was not affected by the treatment allocation. We also found a 90-day mortality of 47%, which is within the range of mortalities recently reported by the REVA Network and the COVID-ICU investigators^25^. In this large epidemiological study, mortality varied between 30% and 50% according to ARDS severity.

Although the hospital length of stay was not affected by the experimental treatment, there was a trend to a shorter time in the ICU in the high-dose group (P =.137). While this could be explained by chance, the finding might be related to a briefer requirement of ventilatory support in patients treated with high-dose dexamethasone. One interesting finding of our trial is that 16% of our patients were still hospitalized either in the primary hospital or in a chronic care/rehabilitation facility 90 days after inclusion.

We found a significant burden of safety issues potentially related to corticosteroids, including nosocomial infections, hyperglycemia, muscle weakness, and delirium. The occurrence of these problems seemed not to be increased by the high-dose treatment. However, this should be interpreted with caution as our trial was not powered to detect minor differences in these safety outcomes.

This study has many limitations. First, it is an open-label study. A double-blind design would be desirable but, given the urgent need for evidence required by the pandemic, we were unable to do it differently. Nevertheless, we believe that our data may provide some useful and interesting findings. Second, the early and unplanned termination of the study due to poor accrual after nine months indicates a failure in our trial process. The reasons for this premature termination were related to the fast-changing dynamics of the pandemic and were not anticipated by us during the trial design. The smaller size probably reduced the power of our study to detect differences in the VFD and other secondary outcomes. Third, the lack of shared procedures for liberation from mechanical ventilation in this multicenter open-label trial produces a potential bias. However, international guidelines commonly used were suggested. Finally, the investigators reported the results and conducted the analysis. Nevertheless, to prevent bias, the analysis was conducted as planned at the writing of the protocol, following such procedures after the termination of the data recollection.

## CONCLUSIONS

This open-label multicenter clinical trial did not show difference in ventilator-free days between high- and low-dose dexamethasone in patients with C-ARDS. However, suggests that a 10-day course of high-dose intravenous dexamethasone improved the time required to liberate these patients from the ventilator compared with the recommend low-dose treatment. This was not associated with a benefit in 90-day mortality or a higher frequency of safety issues. The reduction in the time of mechanical ventilation, even in the absence of a demonstrated treatment effect on mortality, might be critical considering the shortage of ICU resources reported worldwide.

## Data Availability

De-identified data will be available upon request

## Notes

<u>Funding / Support</u>: This research did not receive any specific grant from funding agencies in the public, commercial, or not-for-profit sectors.

<u>Conflicts of interest</u>: None

### Competing Interest Statement

The authors have declared no competing interest.

### Clinical Trial

NCT04395105

### Funding Statement

This research did not receive any specific grant from funding agencies in the public, commercial, or not-for-profit sectors

### Author Declarations

The study was approved at the Research Ethics Committee of CEMIC (Buenos Aires, Argentina) and by the Argentine Society of Intensive Care Medicine (SATI) Ethics Committee

xI have followed all appropriate research reporting guidelines and uploaded the relevant EQUATOR Network research reporting checklist(s) and other pertinent material as supplementary files, if applicable.

