## Supplemental material for "High- Versus Low-Dose Dexamethasone for the Treatment of COVID-19-related Acute Respiratory Distress Syndrome: A Multicenter and Randomized Open-label Clinical Trial"

### ^1^Intensive Care Unit, Hospital Universitario Sede Pombo (Instituto Universitario CEMIC, Centro de Educación Médica e Investigaciones Clínicas), Buenos Aires, Argentina; ^2^Pulmonary Section, CEMIC, Buenos Aires, Argentina; ^3^Intensive Care Unit, Hospital Universitario Sede Saavedra (Instituto Universitario CEMIC), Buenos Aires, Argentina; ^4^Intensive Care Unit, Sanatorio Sagrado Corazón, Buenos Aires, Argentina; ^5^Intensive Care Unit, Clínica Bazterrica, Buenos Aires, Argentina

### Corresponding Author:

Pablo O. Rodriguez

Unidad de Terapia Intensiva, Hospital Universitario Sede Pombo

CEMIC (Centro de Educación Médica e Investigaciones Clínicas)

Av. Cnel. Diaz 2423, Zip Code: 1425, Ciudad Autónoma de Buenos Aires, Argentina

**Content**

### **e-Methods**

#### **Additional information on exclusion criteria**

We enrolled critically ill patients with acute respiratory distress syndrome (ARDS) due to confirmed COVID-19 admitted to the ICU.

The exclusion criteria were the following:

- Pregnant or breastfeeding women,
- Terminal disease (advanced cancer; being under palliative care; cardiovascular, respiratory, or renal disease with a life expectancy less ≤ 1 year),
- Therapeutic limitation (advance directives or do not resuscitate order),
- Severe immunosuppression (non-controlled HIV infection, long-term use of immunosuppressive agents, active cancer),
- Chronic treatment with glucocorticoids for other diseases (≥8 mg prednisone or equivalent),
- Participation in another randomized clinical trial,
- Utilization of low-dose dexamethasone for more than five days due to COVID-19, and
- Consent refusal.

#### **Additional information on clinical and laboratory data**

We collected information in four domains, namely, rate of infections, glycemic control, muscle dysfunction, and delirium, as safety measurements. We also registered the number of microbiologically confirmed infections (ventilator-associated pneumonia, primary bacteremia or catheter-related bloodstream infection, or urinary tract infection). The glycemic control was quantified by peak glucose levels per day and daily cumulative insulin doses, and the muscular strength was daily assessed with the MRC (Medical Research Council) score upon awakening.^1^ This score evaluates the strength from six muscle groups and goes from 0 (complete paralysis) to 60 (normal strength). Finally, the delirium occurrence was evaluated with the CAM-ICU assessment,^2^ and delirium was also defined when neuroleptics were required.

### **e-Results**

#### **Additional information on safety measurements**

Microbiologically confirmed infections were diagnosed in 72 (73%) patients during the first 28 days following randomization. Overall, 150 nosocomial infections were observed during this period. The most frequent diagnoses were ventilator-associated pneumonia (62 [41%]), followed by bacteremia (54 [36%]) and urinary tract infection (24 [16%]). The time from randomization to the first nosocomial infection was 6 (3–8) days. The estimated rate of infections of the control arm was 2.42 (95% CI: 1.92 to 3) per patient per month, and the incidence rate ratio of the high dose group was 1.12 (95% CI: 0.81 to 1.54, P = .502).

According to the mixed-effects linear regression model, the peak finger-prick glucose was 228 (95% CI: 210 to 246) mg/dL during the first day in the control arm. There was not a noticeable change in the 10 days of the study, and the interaction between the time and the experimental treatment was not statistically significant (P for interaction = .977). The insulin dose during the first day of treatment was 28 (95% CI: 14 to 41) IU. Daily doses remained high during the next 10 days, and there was not a significant interaction with the treatment (interaction term P = .549).

Muscle strength could be evaluated in 51 patients, and the minimal MRC score (normal value is 60) recorded per patient was not affected by the treatment allocation: 30 (24–33) in the high-dose versus 28 (24–35) in the low-dose dexamethasone group (P = .954).

Delirium was noted in 28 (57%) patients in the high-dose dexamethasone group. Similarly, this was observed in 27 (55%) cases from the other group (P > .999).

### **e-Figures**

#### **e-Figure 1: Cumulative number of screened and recruited patients.**

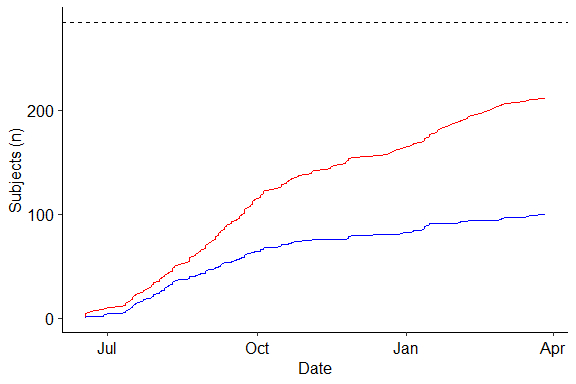

The red and the blue lines correspond to the screened and randomized patients, and the dotted line represents the target number of patients.

#### **e-Figure 2: Expected time for completing the trial according to the recruitment per month.**

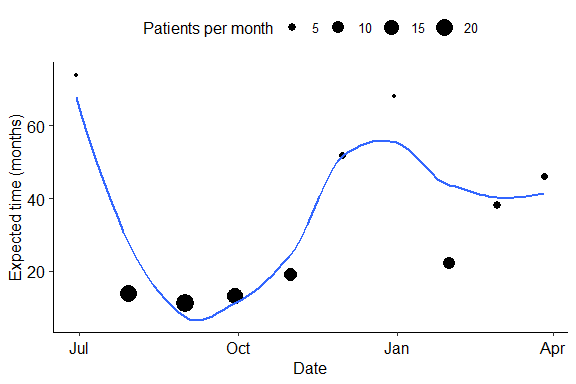

The points represent the expected time in months to achieve the target sample size (n = 284) based on the observed recruitment rate of the previous month. The point size is scaled to the number of subjects effectively recruited per month. The blue line represents a smoothing line using the LOESS method (local polynomial regression).

#### **e-Figure 3: Probability of 90-day survival according to the treatment allocation**

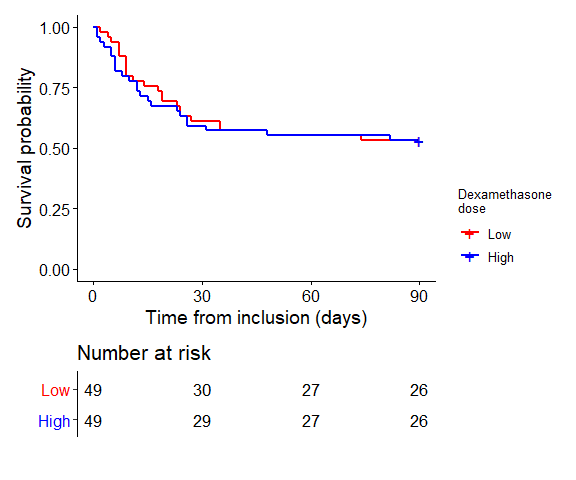

The log-rank test p was 0.862.

### **e-Tables**

#### **e-Table 1: Additional baseline biological data**

|  | **Low-dose dexamethasone** | **High-dose**  **dexamethasone** | **All**  **patients** |
| --- | --- | --- | --- |
|  | *n=51* | *n=49* | *n=100* |
| Hemoglobin (g/dL) | 12.6 (11.45–13.45) | 12.65 (11.38–13.43) | 12.6 (11.4–13.45 |
| White blood cell count (/mm^3^) | 11000 (7785–14125) | 10980 (7847.5–14340) | 11000 (7785–14460) |
| Lymphocyte count (/mm^3^) | 652 (445–935) | 601 (478–798.5) | 644 (453.25–855) |
| ESR (mm/hr) | 76.24±37.38 | 65.07±35.27 | 71±36.26 |
| C-reactive protein (mg/dL) | 11 (6.3–15.5) | 8.29 (6.7–20.17) | 9 (6.5–17.9) |
| Ferritin (ng/mL) | 1420 (644–2357) | 1189.5 (840.12–1917.75) | 1279 (735–2150) |
| Platelet count (**×**1000/mm^3^) | 283 (185.5–351.25) | 236 (183–3015) | 256 (184–320) |
| Thrombin time (%) | 88 (81–100) | 90.5 (79–100) | 89 (80–100) |
| KPTT (seconds) | 33 (31–38) | 34 (32.25–37) | 34 (31–37.5) |
| Fibrinogen (mg/dL) | 646.14±124.85 | 585.2±211.18 | 620.75±160.33 |
| D-Dimer (mcg/mL) | 1.03 (0.38–4.03) | 0.86 (0.41–2.39) | 0.9 (0.39–3.82) |
| Troponin-T (ng/L) | 13.5 (7.32–67) | 10.5 (6.63–24.25) | 11.22 (6.78–44.5) |
| NT-proBNP (pg/mL) | 367 (101.75–1126.25) | 593 (187.1–788) | 468 (146.5–870.5) |
| Lactic acid (mmol/L) | 1.7 (1.4–2.28) | 2.1 (1.55–2.6) | 1.8 (1.4–2.42) |
| Lactic dehydrogenase (IU/L) | 746 (550.5–976) | 668.5 (624.25–760) | 680 (567–908.5) |

#### **e-Table 2: Additional baseline pharmacological treatments and glycemic control**

|  | **Low-dose**  **dexamethasone** | **High-dose**  **dexamethasone** | **All**  **patients** |
| --- | --- | --- | --- |
|  | *n=51* | *n=49* | *n=100* |
| Previous dexamethasone (days) | 2 (1–4.25) | 2 (1–4) | 2 (1–4) |
| Antibiotics (%) | 41 (80.39%) | 39 (79.59%) | 80 (80%) |
| Macrolides (%) | 20 (39.22%) | 13 (26.53%) | 33 (33%) |
| Oseltamivir (%) | 8 (15.69%) | 4 (8.16%) | 12 (12%) |
| Fluid input (mL) | 2119 (1700–3052) | 2442 (1775.25–3090) | 2381.5 (1718.5–3080.5) |
| Fluid output (mL) | 1300 (1000–1720) | 1200 (850–1750) | 1250 (960–1742.5) |
| Minimum glycemia (mg/dL) | 147 (126–170) | 139 (112–151) | 140 (120–166) |
| Maximum glycemia (mg/dL) | 193 (150.25–256.5) | 188 (160.5–239) | 189 (156–243) |
| Insulin (IU/day) | 0 (0–14.5) | 0 (0–6) | 0 (0–10) |
